# Availability of benign missense variant “truthsets” for validation of functional assays: current status and a novel systematic approach

**DOI:** 10.1101/2025.03.20.25324299

**Authors:** Charlie F. Rowlands, Sophie Allen, Alice Garrett, Miranda Durkie, George J. Burghel, Rachel Robinson, Alison Callaway, Joanne Field, Bethan Frugtniet, Sheila Palmer-Smith, Jonathan Grant, Judith Pagan, Trudi McDevitt, Katie Snape, Helen Hanson, Terri McVeigh, Clare Turnbull, CanVIG-UK

## Abstract

Multiplex assays of variant effect (MAVEs) provide promising new sources of functional evidence, potentially empowering improved classification of germline genomic variants, particularly rare missense variants, which are commonly assigned as VUS (variants of uncertain significance). However, paradoxically, quantification of clinically applicable evidence strengths for MAVEs requires construction of “truthsets” comprising missense variants already robustly classified as pathogenic and benign. In this study, we demonstrate how benign truthset size is the primary driver of applicable functional evidence towards pathogenicity (PS3). We demonstrate, when using existing ClinVar classifications as a source of benign missense truthset variants, for only 19.8% (23/116) of established cancer susceptibility genes was a PS3 evidence strength of “strong” attainable when simulating validation for a hypothetical new MAVE (applying also favourable assumption of perfect concordance). We describe a “proactive-systematic” framework for benign truthset construction, in which all possible missense variants in a gene of interest are concurrently assessed for assignation of (likely) benignity via established ACMG/AMP combination rules including population frequency, *in silico* evidence codes and case-control signal. We apply this framework to eight hereditary breast and ovarian cancer genes, demonstrating that proactive-systematically generated benign missense truthsets allow maximum application of PS3 at greater (or equivalent) strength – reaching “moderate” for *CHEK2* and “strong” for the other seven genes – than those derived from ClinVar ≥2* classifications alone. We propose, given many genes have few existing benign-classified missense variants, application of this proactive-systematic framework to disease genes more broadly will be important for leveraging full value from MAVEs.

## Introduction

With rapid advances in high-throughput molecular and bioinformatic pipelines for genomic sequencing, the clinical interpretation of detected variants has become the primary bottleneck in clinical genetic testing^1^. Most commonly problematic for interpretation of pathogenicity are rare missense variants, which are in aggregate frequent^2^. For the majority of such missense variants encountered on clinical testing, clinical case data are unavailable or insufficient to provide informative signals. and they are ascribed as variants of uncertain significance (VUS)^2–5^.

One attractive source of information to assist in resolving VUS classifications, and to determine if a variant may be deleterious or of neutral impact, are functional assays of variant effect. Recently, through concurrent advances in high throughput sequencing and gene-editing technologies, multiplexed assays of variant effect (MAVEs) have provided systematic data on the functional impact of many thousands of variants^4,6,7^. Such assays offer potential pre-annotation of (nearly) every possible variant that could arise ahead of it being observed clinically^8–10^. However, for either low- or high-throughput assays, an assay readout of neutral or deleterious function should not be automatically presumed to recapitulate clinical pathogenicity without suitably robust validation.

In 2019, on behalf of the ClinGen SVI group, Brnich *et al*. described a methodology to quantify the PS3/BS3 functional evidence criteria, such that assays could be assigned individualised evidence strengths based on their ability to discriminate between two “truthsets” of pathogenic and benign variants^11^. This specification by Brnich *et al*. built upon the looser stipulation in the 2015 ACMG-AMP variant interpretation framework in which assignation of evidence towards pathogenicity (PS3) or benignity (BS3) was allowed where “well-established *in vitro* or *in vivo* studies [are] supportive of/[show] no damaging effect on the gene or gene product”^12^. The Brnich *et al.* methodology incorporated the subsequent evolution of the 2015 ACMG-AMP guidelines by Tavtigian *et al.*, which provided a Bayesian framework by which evidence items may be assigned strengths dependent on the likelihood ratio (LR) afforded towards pathogenicity or benignity^13^.

Missense variants may pose particular challenges in assessment and application of functional evidence, as the mechanism of impact of missense variants on protein function may vary, with effects for both pathogenic and benign variants potentially more likely to lie towards the intermediate zone of assay dynamic range^14^. The complex spectrum of missense variant effects is thus not necessarily well-reflected in standard preliminary calibration/validation of assay performance (for loss-of-function genes) against nonsense/frameshift and synonymous variants. It is thus argued that, as a second phase of validation, assays should be clinically validated against truthsets of benign-classified and pathogenic-classified missense variants (in particular given that missense variants are the group for which functional data will more strongly drive a classification of pathogenicity)^15^.

Publicly available databases, most commonly ClinVar, have typically served as the *de facto* sources of truthset variants^3^. However, most missense variants in ClinVar are classified as VUS^4^. Furthermore, for many missense variants in ClinVar classified as (likely) benign (B/LB) or (likely) pathogenic (P/LP), the level of detailing of evidence items underpinning these classifications may vary substantially. Accordingly, it may be unclear for many variants whether their extant classifications have been attained through inclusion of functional data (PS3/BS3 codes). This circularity renders problematic inclusion of such variants in truthsets for evaluating the evidence strength to be assigned for a new assay^16^.

Limited availability of robust missense truthset variants is a primary roadblock in regard of our ability to best leverage for clinical variant interpretation the recent surge in availability of data from MAVEs^15^. This will be particularly problematic for understudied genes and genes that are rarer causes of disease (which, paradoxically, constitute the genes for which variant interpretation stands to gain most from integration of functional evidence).

In recognition of the incipient emergence into the clinical diagnostic arena of many new MAVEs, we sought to better evaluate our ‘preparedness’ in regard of missense truthsets for validation. In the work presented here, we:

- Demonstrate that the number of benign-classified truthset variants is (paradoxically) the principal driver of the strength with which the PS3 code towards pathogenicity can be assigned.
- Examine the availability from ClinVar of truthset variants for 116 cancer susceptibility genes that are routinely tested for in clinical practice, considering in detail eight genes associated with hereditary breast and ovarian cancer (HBOC).
- Describe a “proactive-systematic” approach to construction of benign missense truthsets based on the original ACMG-AMP combination rules. We demonstrate, for the eight HBOC genes, the equal or improved power for MAVE assay validation and accordant strength for allocation of PS3 afforded through this proactive-systematic approach to construction of benign missense truthsets.

## Methods

### Extraction of ClinVar classifications and comments

Counts of ClinVar missense variant classifications were retrieved for eight HBOC genes of interest (*BRCA1* [MIM: 113705], *BRCA2* [MIM: 600185], *PALB2* [MIM: 610355]*, ATM* [MIM: 607585], *CHEK2* [MIM: 604373], *RAD51C* [MIM: 602774], *RAD51D* [MIM: 602954] and *BRIP1* [MIM: 605882]) and 108 additional inherited cancer susceptibility genes through direct querying of the ClinVar bulk data file download, and attached comments collated for HBOC gene classifications via the ClinVar API.

### Extraction of gnomAD, UK Biobank and BRIDGES population variant data

The coding sequence of the MANE transcript for each HBOC gene was extracted using Ensembl BioMart^17^ (v111) and bespoke Python scripts used to generate all possible SNVs in these transcripts. Variants overlapping these coding regions were extracted and processed from the UK Biobank (UKB) final exome release population VCF^18^ and the gnomAD v2.1.1^19^ variant count bulk file download using the bcftools view command (see Supplemental Methods).

Collated variant counts were stratified by non-founder ancestry grouping (in gnomAD and UKB) and case vs. control status (in UKB, see Supplemental Methods for ancestry and case cohort definitions). Counts of missense variant observations in the BRIDGES unselected breast cancer case and control series were also retrieved from the primary publication^20^.

### Assignation of putative pathogenicity and benignity flags

We devised a “proactive-systematic” approach to construction of benign truthsets based on *in silico* and population frequency ACMG/AMP evidence codes. All possible missense variants in each HBOC gene were annotated with BayesDel^21^, REVEL^22^ and SpliceAI^23^ *in silico* scores using their respective flat file data downloads, ensuring matching of transcripts where scores were transcript-specific. Variants were excluded from proactive-systematically generated benign truthsets if *in silico* predictions indicated possible pathogenicity (BayesDel score ≥0.13 or REVEL score ≥0.644, as per previously calibrated PP3_supporting thresholds^24^) or if their maximum SpliceAI score was ≥0.2.

We further excluded from benign truthsets any missense variant with observed effect size (i.e. association with gene-associated cancer type) of odds ratio (OR) >2 and lower confidence interval (CI) limit >1 in any sub-ancestry in the BRIDGES and UKB datasets, as described above and in Supplemental Methods.

Remaining missense variants with REVEL scores ≤0.29 were considered to have met the BP4_sup criterion as per the aforementioned thresholds.^24^

Population frequency thresholds for application of BA1 and BS1 were identified via calculation of gene-phenotype dyad-specific maximum tolerated allele frequencies (MTAFs; Supplemental Methods), as previously described^25^, as well as BS1_supporting (BS1_sup) thresholds, as employed in several ClinGen-ratified VCEP guidelines^26–29^. Variants present at frequencies above these thresholds in any superpopulation or non-founder subpopulation in gnomAD or UKB were annotated with the corresponding population evidence code.

Proactive-systematically generated benign truthsets of varying stringency were constructed according to different combinations of evidence codes, as detailed in the Results, Figure S1 and Table S1.

## Results

### Impact of truthset size on applicable PS3/BS3 evidence strength

We first investigated the impact of varying pathogenic/benign truthset size on the applicable PS3/BS3 strengths of assays using the established Bayesian framework developed by Brnich *et al*.^11^ (Table 1). We observed that, holding the pathogenic truthset size constant for a hypothetical perfectly concordant assay (i.e. one exhibiting complete consistency between assay readout and truthset classification), an increase in benign truthset size led to an accordant increase in applicable exponent points (EPs) for the PS3 code, and thereby in applicable PS3 strength (Table 1A). Conversely, holding benign truthset size constant and increasing pathogenic truthset size led to an increase in the applicable BS3 strength, illustrating the inverse relationship between the two.

**Table 1.** Quantification of impact of truthset size on applicable evidence strengths under the Brnich framework for PS3/BS3 strength evaluation. Displayed are the assay strengths in terms of both exponent points (EPs) and equivalent 2015 ACMG/AMP evidence strengths for hypothetical MAVEs exhibiting (A) perfect discrimination between and varying levels of discordance with assay readout of (B) pathogenic, (C) benign and (D) both types of truthset variant in truthsets of varying sizes. For perfectly discriminant assays, PS3 strength is entirely dependent on benign truthset size, and BS3 strength entirely on pathogenic truthset size. In the presence of any discordance with assay readout, benign and pathogenic truthset size remain the respective drivers of PS3 and BS3 strength, but with the opposing truthset (i.e. the pathogenic truthset for PS3 and the benign for BS3) having some mild influence on applicable EPs. SUP, supporting; MOD, moderate; STR, strong

| Number of truthset variants |  | # truthset variants by assay readout |  |  |  | ACMG EPs |  |
| --- | --- | --- | --- | --- | --- | --- | --- |
| Pathogenic | Benign | Pathogenic readout |  | Benign readout |  | PS3 | BS3 |
|  |  | Path. | Ben. | Path. | Ben. |  |  |
| (A) Assay displaying perfect concordance with truthsets |  |  |  |  |  |  |  |
| 4 | 4 | 4 | 0 | 0 | 4 | 1.89 (SUP) | 1.89 (SUP) |
| 10 |  | 10 | 0 | 0 | 4 | 1.89 (SUP) | 3.14 (MOD) |
| 100 |  | 100 | 0 | 0 | 4 | 1.89 (SUP) | 6.29 (STR) |
| 4 | 10 | 4 | 0 | 0 | 10 | 3.14 (MOD) | 1.89 (SUP) |
| 10 |  | 10 | 0 | 0 | 10 | 3.14 (MOD) | 3.14 (MOD) |
| 100 |  | 100 | 0 | 0 | 10 | 3.14 (MOD) | 6.29 (STR) |
| 4 | 100 | 4 | 0 | 0 | 100 | 6.29 (STR) | 1.89 (SUP) |
| 10 |  | 10 | 0 | 0 | 100 | 6.29 (STR) | 3.14 (MOD) |
| 100 |  | 100 | 0 | 0 | 100 | 6.29 (STR) | 6.29 (STR) |
| (B) Assay exhibiting discordance for variants in benign truthset |  |  |  |  |  |  |  |
| 4 | 4 | 4 | 1 | 0 | 3 | 0.95 (None) | 1.50 (SUP) |
| 10 |  | 10 | 1 | 0 | 3 | 0.95 (None) | 2.75 (MOD) |
| 100 |  | 100 | 1 | 0 | 3 | 0.95 (None) | 5.90 (STR) |
| 4 | 10 | 4 | 2 | 0 | 8 | 1.64 (SUP) | 1.59 (SUP) |
| 10 |  | 10 | 2 | 0 | 8 | 1.64 (SUP) | 2.84 (MOD) |
| 100 |  | 100 | 2 | 0 | 8 | 1.64 (SUP) | 5.98 (STR) |
| 4 | 100 | 4 | 5 | 0 | 95 | 3.84 (MOD) | 1.82 (SUP) |
| 10 |  | 10 | 5 | 0 | 95 | 3.84 (MOD) | 3.07 (MOD) |
| 100 |  | 100 | 5 | 0 | 95 | 3.84 (MOD) | b.22 STR |
| (C) Assay exhibiting discordance for variants in pathogenic truthset |  |  |  |  |  |  |  |
| 4 | 4 | 3 | 0 | 1 | 4 | 1.50 (SUP) | 0.95 (None) |
| 10 |  | 8 | 0 | 2 | 4 | 1.59 (SUP) | 1.64 (SUP) |
| 100 |  | 95 | 0 | 5 | 4 | 1.82 (SUP) | 3.84 (MOD) |
| 4 | 10 | 3 | 0 | 1 | 10 | 2.75 (MOD) | 0.95 (None) |
| 10 |  | 8 | 0 | 2 | 10 | 2.84 (MOD) | 1.64 (SUP) |
| 100 |  | 95 | 0 | 5 | 10 | 3.07 (MOD) | 3.84 (MOD) |
| 4 | 100 | 3 | 0 | 1 | 100 | 5.90 (STR) | 0.95 (None) |
| 10 |  | 8 | 0 | 2 | 100 | 5.98 (STR) | 1.64 (SUP) |
| 100 |  | 95 | 0 | 5 | 100 | 6.22 (STR) | 3.84MOD |
| (D) Assay exhibiting discordance for variants in both pathogenic and benign truthsets |  |  |  |  |  |  |  |
| 4 | 4 | 3 | 1 | 1 | 3 | 0.55 (None) | 0.55 (None) |
| 10 |  | 8 | 1 | 2 | 3 | 0.64 (None) | 1.25 (SUP) |
| 100 |  | 95 | 1 | 5 | 3 | 0.88 (None) | 3.45 (MOD) |
| 4 | 10 | 3 | 2 | 1 | 8 | 1.25 (SUP) | 0.64 (None) |
| 10 |  | 8 | 2 | 2 | 8 | 1.34 (SUP) | 1.34 (SUP) |
| 100 |  | 95 | 2 | 5 | 8 | 1.57 (SUP) | 3.54 (MOD) |
| 4 | 100 | 3 | 5 | 1 | 95 | 3.45 (MOD) | 0.88 (None) |
| 10 |  | 8 | 5 | 2 | 95 | 3.54 (MOD) | 1.57 (SUP) |
| 100 |  | 95 | 5 | 5 | 95 | 3.77 (MOD) | 3.77 (MOD) |

PS3 strength is also wholly determined by the benign truthset size when discordance between the assay readouts and truthset classifications is restricted to deleterious readouts for benign-classified truthset variants (Table 1B). Likewise, applicable BS3 strength is wholly driven by the size of the pathogenic truthset when discordancy is restricted to neutral readouts for pathogenic-classified truthset variants (Table 1C).

### Current availability of benign and pathogenic missense variant truthsets for cancer susceptibility genes

We next sought to quantify the number of missense variants available with existing classifications of (i) B/LB and (ii) P/LP in ClinVar (Table 2). We first examined eight widely-tested genes implicated in hereditary breast and ovarian cancer (HBOC), namely *BRCA1*, *BRCA2*, *PALB2*, *ATM*, *CHEK2*, *RAD51C*, *RAD51D* and *BRIP1*.

**Table 2.** Comparison of the number of missense variant ClinVar classifications for eight HBOC genes. Shown are the counts of variants (of any variant type and of missense variants alone) with ClinVar classifications of (likely) pathogenic and (likely) benign for eight selected hereditary breast and ovarian cancer (HBOC) genes. Counts are additionally stratified by review status (≥2* or ≥1*). Across all genes and review status thresholds, it was observed that missense variants constituted a minority of total (likely) benign/pathogenic classifications, disproportionate to their frequency in clinical testing. This imbalance was particularly noticeable for *CHEK2*, *RAD51C*, *RAD51D* and *BRIP1*. B, benign; LB, likely benign; LP, likely pathogenic; P, pathogenic.

| Gene | ClinVar review status | # ClinVar-classified variants |  |  |  |  |  |  |  |  |  |  |  |  |  |
| --- | --- | --- | --- | --- | --- | --- | --- | --- | --- | --- | --- | --- | --- | --- | --- |
|  |  | P/LP |  |  |  |  |  |  | B/LB |  |  |  |  |  |  |
|  |  | All variants |  |  | Missense variants |  |  | % missense | All variants |  |  | Missense variants |  |  | % missense |
|  |  | P | LP | P+LP | P | LP | P+LP |  | B | LB | B+LB | B | LB | B+LB |  |
| Inherited breast cancer susceptibility |  |  |  |  |  |  |  |  |  |  |  |  |  |  |  |
| BRCA1 | ≥2* | 2641 | 31 | 2672 | 90 | 18 | 108 | 4.0% | 746 | 1028 | 1774 | 152 | 31 | 183 | 10.3% |
|  | ≥1* | 3358 | 175 | 3533 | 109 | 63 | 172 | 4.9% | 789 | 2384 | 3173 | 160 | 142 | 302 | 9.5% |
| BRCA2 | ≥2* | 3310 | 25 | 3335 | 41 | 7 | 48 | 1.4% | 790 | 1761 | 2551 | 153 | 48 | 201 | 7.9% |
|  | ≥1* | 4488 | 260 | 4748 | 45 | 24 | 69 | 1.5% | 821 | 3572 | 4393 | 157 | 163 | 320 | 7.3% |
| PALB2 | ≥2* | 624 | 39 | 663 | 1 | 0 | 1 | 0.2% | 87 | 502 | 589 | 11 | 0 | 11 | 1.9% |
|  | ≥1* | 1098 | 121 | 1219 | 1 | 1 | 2 | 0.2% | 114 | 1165 | 1279 | 12 | 13 | 25 | 2.0% |
| ATM | ≥2* | 1057 | 131 | 1188 | 24 | 15 | 39 | 3.3% | 244 | 1601 | 1845 | 25 | 0 | 25 | 1.4% |
|  | ≥1* | 2270 | 469 | 2739 | 29 | 30 | 59 | 2.2% | 374 | 4136 | 4510 | 31 | 20 | 51 | 1.1% |
| CHEK2 | ≥2* | 287 | 68 | 355 | 3 | 1 | 4 | 1.1% | 36 | 290 | 326 | 1 | 0 | 1 | 0.3% |
|  | ≥1* | 658 | 187 | 845 | 3 | 3 | 6 | 0.7% | 72 | 799 | 871 | 1 | 4 | 5 | 0.6% |
| Inherited ovarian cancer susceptibility |  |  |  |  |  |  |  |  |  |  |  |  |  |  |  |
| RAD51C | ≥2* | 104 | 19 | 123 | 5 | 2 | 7 | 5.7% | 26 | 193 | 219 | 2 | 0 | 2 | 0.9% |
|  | ≥1* | 181 | 61 | 242 | 5 | 2 | 7 | 2.9% | 35 | 486 | 521 | 3 | 1 | 4 | 0.8% |
| RAD51D | ≥2* | 49 | 22 | 71 | 0 | 0 | 0 | 0% | 25 | 198 | 223 | 3 | 0 | 3 | 1.3% |
|  | ≥1* | 119 | 70 | 189 | 0 | 0 | 0 | 0% | 40 | 510 | 550 | 3 | 1 | 4 | 0.7% |
| BRIP1 | ≥2* | 318 | 64 | 382 | 2 | 1 | 3 | 0.8% | 69 | 541 | 610 | 5 | 0 | 5 | 0.8% |
|  | ≥1* | 613 | 172 | 785 | 2 | 4 | 6 | 0.8% | 110 | 1339 | 1449 | 5 | 18 | 24 | 1.7% |

We first quantified the relative deposition of missense variant classifications in ClinVar for these eight HBOC genes (Table 2). What was first notable was the modest absolute numbers of P/LP and B/LB classifications for missense variants in these widely-tested genes: the median average across the genes of 1.28/1.11% of P/LP and 1.39/1.35% of B/LB ≥1*/≥2* classifications being missense variants. Available B/LB classifications of ≥1*/≥2* varied widely between genes (Table 2) but were strikingly low for *CHEK2* (5/1), *RAD51C* (4/2) and *RAD51D* (4/3). By gene, of ≥1* B/LB missense variants, a median of 51.0% were “benign” (versus “likely benign”) whilst a median of 50.0% were ≥2* (versus ≥1*). These data highlight the disproportionate paucity in ClinVar of (i) missense variants and (ii) high-confidence classifications of benignity therein for many genes.

We next evaluated the hypothetical evidence strengths applicable for MAVEs if using ClinVar as a source of missense variants for construction of “benignity truthsets”. Considering in the first instance a stringent inclusion criterion for prospective benign missense truthset variants of B/LB with ≥2* ClinVar review status, for perfectly concordant MAVEs, PS3 would be applicable at a maximum of “strong” for assays of *ATM*, *BRCA1* and *BRCA2*, at “moderate” for *PALB2* and *BRIP1*, and would not be applicable at all for *CHEK2, RAD51C* or *RAD51D*. The PS3 strengths attainable for MAVEs in these genes increase somewhat on inclusion of ClinVar B/LB variants with only 1* review status (classifications based on a single submitter, Table 2), with moderate/supporting strength now hypothetically attainable for *CHEK2, RAD51C* and *RAD51D*.

We extended this analysis to 116 established cancer susceptibility genes (CSGs) with which are routinely tested for clinically (and are included in the NHS National Test Directory; Table 3). For a perfectly concordant assay, the minimum number of benign truthset variants permitting application of PS3 at strong is 19. Sufficient ClinVar B/LB missense classifications to meet this threshold were available for only 19.8% (≥2*; 23/116) and 36.2% (≥1*; 42/116) of these genes (Figure S2). These findings highlight the relative paucity of ClinVar B/LB missense classifications in this broader group of clinically-tested CSGs.

**Table 3.** Comparison of numbers of pathogenic/likely pathogenic and benign/likely benign ClinVar classifications for missense variants in 116 selected inherited cancer susceptibility genes. Relative numbers of ClinVar missense classifications were counted for each gene from a download of the ClinVar bulk data file.

| Gene | # ClinVar classifications for missense variants |  |  |  |
| --- | --- | --- | --- | --- |
|  | P/LP |  | B/LB |  |
|  | ≥1* | ≥2* | ≥1* | ≥2* |
| AIP | 2 | 1 | 2 | 2 |
| ALK | 5 | 1 | 36 | 27 |
| ANKRD26 | 0 | 0 | 37 | 24 |
| APC | 17 | 12 | 59 | 35 |
| ATM | 59 | 39 | 51 | 25 |
| BAP1 | 8 | 0 | 6 | 6 |
| BARD1 | 2 | 1 | 15 | 12 |
| BLM | 4 | 2 | 29 | 11 |
| BMPR1A | 15 | 2 | 1 | 1 |
| BRCA1 | 172 | 108 | 302 | 183 |
| BRCA2 | 69 | 48 | 320 | 201 |
| BRIP1 | 6 | 3 | 24 | 5 |
| BUB1B | 3 | 1 | 19 | 8 |
| CBL | 12 | 6 | 19 | 11 |
| CDC73 | 2 | 0 | 1 | 0 |
| CDH1 | 12 | 7 | 81 | 75 |
| CDK4 | 2 | 2 | 2 | 0 |
| CDKN1C | 0 | 0 | 0 | 0 |
| CDKN2A | 36 | 25 | 4 | 3 |
| CEBPA | 2 | 0 | 3 | 0 |
| CEP57 | 0 | 0 | 7 | 2 |
| CHEK2 | 6 | 4 | 5 | 1 |
| CYLD | 2 | 0 | 6 | 2 |
| DDB2 | 1 | 0 | 4 | 1 |
| DDX41 | 6 | 3 | 1 | 0 |
| DICER1 | 40 | 13 | 28 | 25 |
| DIS3L2 | 1 | 0 | 10 | 3 |
| EGFR | 6 | 0 | 13 | 8 |
| EPCAM | 0 | 0 | 17 | 9 |
| ERCC2 | 19 | 14 | 7 | 5 |
| ERCC3 | 1 | 0 | 6 | 1 |
| ERCC4 | 5 | 2 | 8 | 8 |
| ERCC5 | 5 | 1 | 20 | 15 |
| ETV6 | 8 | 5 | 4 | 3 |
| EXT1 | 36 | 12 | 12 | 5 |
| EXT2 | 13 | 1 | 18 | 6 |
| EZH2 | 28 | 4 | 23 | 2 |
| FANCA | 40 | 22 | 57 | 30 |
| FANCB | 1 | 0 | 49 | 20 |
| FANCC | 3 | 1 | 14 | 4 |
| FANCD2 | 6 | 2 | 18 | 15 |
| FANCE | 1 | 1 | 8 | 5 |
| FANCF | 0 | 0 | 5 | 3 |
| FANCG | 5 | 2 | 7 | 5 |
| FANCI | 2 | 1 | 21 | 17 |
| FANCL | 0 | 0 | 7 | 5 |
| FANCM | 0 | 0 | 19 | 18 |
| FH | 134 | 53 | 7 | 5 |
| FLCN | 5 | 1 | 2 | 3 |
| GATA1 | 9 | 1 | 36 | 4 |
| GATA2 | 0 | 0 | 37 | 0 |
| GPC3 | 2 | 12 | 59 | 14 |
| GREM1 | 0 | 39 | 52 | 0 |
| HNF1A | 95 | 0 | 6 | 19 |
| HRAS | 27 | 1 | 15 | 2 |
| KIT | 18 | 2 | 29 | 5 |
| KRAS | 58 | 2 | 1 | 0 |
| LZTR1 | 21 | 108 | 302 | 3 |
|  | P/LP |  | B/LB |  |
|  | ≥1* | ≥2* | ≥1* | ≥2* |
| MAX | 2 | 0 | 0 | 0 |
| MEN1 | 112 | 41 | 10 | 4 |
| MET | 6 | 4 | 17 | 13 |
| MITF | 8 | 2 | 6 | 4 |
| MLH1 | 162 | 120 | 34 | 30 |
| MSH2 | 123 | 89 | 212 | 47 |
| MSH6 | 42 | 29 | 71 | 27 |
| MUTYH | 27 | 21 | 11 | 7 |
| NBN | 2 | 0 | 20 | 7 |
| NF1 | 390 | 170 | 23 | 9 |
| NF2 | 3 | 2 | 5 | 3 |
| NSD1 | 131 | 24 | 147 | 62 |
| NTHL1 | 0 | 0 | 0 | 0 |
| PALB2 | 2 | 1 | 25 | 11 |
| PAX5 | 5 | 0 | 5 | 5 |
| PDGFRA | 1 | 0 | 15 | 11 |
| PHOX2B | 3 | 1 | 3 | 0 |
| PMS2 | 20 | 12 | 28 | 16 |
| POLD1 | 3 | 2 | 15 | 12 |
| POLE | 6 | 1 | 31 | 20 |
| POT1 | 4 | 0 | 10 | 5 |
| PPM1D | 1 | 0 | 25 | 5 |
| PRF1 | 38 | 23 | 3 | 3 |
| PRKAR1A | 5 | 1 | 2 | 0 |
| PTCH1 | 40 | 8 | 163 | 90 |
| PTEN | 191 | 126 | 5 | 5 |
| RAD51C | 7 | 7 | 4 | 2 |
| RAD51D | 0 | 0 | 4 | 3 |
| RB1 | 26 | 9 | 34 | 31 |
| RECQL4 | 6 | 2 | 45 | 30 |
| RET | 60 | 46 | 19 | 8 |
| RHBDF2 | 2 | 0 | 23 | 13 |
| RNF43 | 0 | 0 | 15 | 13 |
| RUNX1 | 0 | 0 | 0 | 0 |
| SBDS | 8 | 3 | 1 | 1 |
| SDHA | 9 | 7 | 5 | 5 |
| SDHAF2 | 1 | 1 | 5 | 4 |
| SDHB | 64 | 41 | 4 | 3 |
| SDHC | 7 | 5 | 1 | 0 |
| SDHD | 21 | 14 | 3 | 2 |
| SLX4 | 0 | 0 | 38 | 32 |
| SMAD4 | 22 | 10 | 4 | 2 |
| SMARCA4 | 49 | 7 | 10 | 7 |
| SMARCB1 | 8 | 5 | 1 | 0 |
| SMARCE1 | 9 | 1 | 1 | 1 |
| SPRED1 | 3 | 2 | 12 | 6 |
| STK11 | 32 | 10 | 4 | 2 |
| SUFU | 0 | 0 | 3 | 1 |
| TERT | 27 | 13 | 31 | 7 |
| TMEM127 | 1 | 0 | 4 | 4 |
| TP53 | 183 | 147 | 106 | 64 |
| TSC1 | 22 | 2 | 99 | 50 |
| TSC2 | 138 | 45 | 244 | 104 |
| VHL | 146 | 87 | 4 | 3 |
| WRN | 4 | 0 | 16 | 12 |
| WT1 | 31 | 9 | 5 | 2 |
| XPA | 2 | 1 | 6 | 6 |
| XPC | 2 | 1 | 19 | 14 |

To avoid bias and circularity, truthsets for evaluation of MAVE strength should not include variants classified using any functional data^16^. For missense variants with B/LB ClinVar classifications in the eight HBOC genes selected above, we further explored the composition of the evidence contributing to their assignation as benign through analysis of the text provided against classifications in ClinVar (Supplemental Methods; Figure S3). We could ascertain explicitly or implicitly for 79.1% (239/302) of *BRCA1* and 81.3% (260/320) of *BRCA2* missense variants classified as B/LB in ClinVar that this classification had been achieved without application of functional evidence codes. However, for the remaining 6 genes, for only 39.8% (45/113) of variants could the B/LB classification be shown explicitly or implicitly to exclude functional evidence, highlighting the limitations of ClinVar as a source of truthset variants for these genes. As might be predicted, for 2* classifications, it was more common to identify at least one contributing classification for which application of functional data could be excluded (whether explicitly or implicitly); 1* (single-submitter) classifications were more likely to be unclear or to omit comments entirely (Figure S3).

### Proactive-systematic generation of benign missense truthsets for eight HBOC genes

#### Workflow for proactive-systematic generation of benign missense truthsets

We thus went on to devise a proactive approach for construction of benign missense truthsets using established ACMG-AMP criteria indicative of benignity, hereafter termed the “proactive-systematic” approach (Figure S1). We firstly applied filters to exclude from benign truthsets any missense variant with evidence of pathogenicity on the basis of existing ClinVar classifications, *in silico* missense or splicing predictions, or evidence of case-control signal of association (see Methods). We then used ancestry subgroup data from both UK Biobank (UKB) and gnomAD v2.1.1 to annotate remaining missense variants with codes relating to population frequency, namely the BA1 stand-alone code, the BS1 code and a BS1_sup code (see Supplemental Methods). Finally, we assigned evidence towards benignity using the BP4 criterion for *in silico* evidence using REVEL thresholds as previously established^24^.

In accordance with the 2015 ACMG/AMP framework rules for evidence combination, we considered as the “standard” designation for variant benignity either (i) a BA1 annotation (with or without any *in silico* evidence), or (ii) annotation of both a BS1/BS1_sup population frequency code *and* a BP4 *in silico* code of any strength. However, in fitting with emergent v4.0 ClinGen-ACMG updates^30^, we also explored inclusion as likely benign based on the more permissive thresholds of a single evidence item towards benignity of (i) strong or greater, and (ii) supporting or greater strength (Table S1).

The distinct BA1, BS1 and BS1_sup allele frequency thresholds were calculated by specifying allelic heterogeneity values of 1, 0.1 and 0.05 during MTAF construction (see Supplemental Methods). Allocation of the population codes (BA1/BS1/BS1_sup) was based on the maximum tolerated allele count (MTAC) for the given population/subpopulation, which is the number of observations reflecting the upper 95% confidence interval for the corresponding MTAF. We explored applying, as an additional level of conservatism, a minimum number of individuals in whom the variant must be observed for application of these population codes of ≥2 or ≥5 (Table S1); this would constitute an additional cautionary measure against stochastic occurrences and/or erroneous genotyping. For the primary results presented, we stipulated observation of ≥2 individuals to attain BS1/BS1_sup and ≥5 to attain BA1.

#### Quantification of benign missense truthset sizes and applicable ACMG evidence strengths

Using the proactive-systematic construction approach under “standard” ACMG/AMP combination rules and requiring ≥2/≥5 variant carriers for application of BS1/BA1 population codes, respectively, we generated benign missense truthsets of between 12 (for *CHEK2*) and 188 (for *BRCA2*) variants (Table 4; Table S2). For all genes except *CHEK2*, these truthsets would be of sufficient size to enable evidence points (EPs) of between ≥4 (4.69 to 7.15) towards pathogenicity, equivalent to application of PS3 at strong (assuming a perfectly concordant assay). For *CHEK2*, the equivalent EPs applicable towards PS3 were 3.39, equating to moderate strength. We noted that, under the rules applied, the count of variant carriers in the given population group whereby a population evidence code was applied ranged from 2-77 for application of BS1_sup and 2-585 for application of BS1. For BA1, the range was 5-177,212, although, aside from four *PALB2* variants identified in the small UKB African and Chinese sub-ancestries, for all other instances in which BA1 was awarded the number of variant carriers was ≥10 (Figure S4).

**Table 4.** Evidence strengths applicable for hypothetical MAVEs in eight HBOC genes using proactive-systematically constructed validation truthsets of benign missense variants. Displayed are the applicable exponent points and corresponding evidence strengths, calculated as previously described_11_ for simulated MAVEs in eight HBOC genes using both ClinVar and our proactive-systematic method as sources of truthsets of benign missense truthsets. For the proactive-systematic approach, we required ≥2 variant carriers for application of BS1/BS1_sup population codes and ≥5 carriers for application of BA1. For both benign truthset construction methodologies, the pathogenic missense truthsets comprise the total number of pathogenic missense variants present in ClinVar, stratified by review status (≥2* or ≥1*); accordingly, the EPs_Ben_ shown for the ClinVar-based analysis are also applicable to the proactive-systematic analysis. Strengths are assigned assuming all variants are covered by the MAVE, and there is perfect discrimination of the truthset variants. *For *RAD51D*, an arbitrary pathogenic truthset size of 1 was used for calculation of PS3 EPs and strength due to an absence of eligible pathogenic missense variants in ClinVar. EPs_Ben_, exponent points towards pathogenicity; EPs_Path_, exponent points towards pathogenicity; MOD, moderate; STR, strong; SUP, supporting.

| Gene | ClinVar review status threshold | Benign missense truthset construction methodology |  |  |  |  |  |
| --- | --- | --- | --- | --- | --- | --- | --- |
|  |  | ClinVar |  |  |  | Proactive-systematic |  |
| | | # (Likely) pathogenic | # (Likely) benign | $EP_{S_{Path}}$ | $EP_{S_{Ben}}$ | # (Likely) benign | $EP_{S_{Path}}$ |
| *BRCA1* | $\geq 2^*$ | 108 | 183 | 7.11 (STR) | 6.39 (STR) | 34 | 4.82 (STR) |
| | $\geq 1^*$ | 172 | 302 | 7.80 (STR) | 7.03 (STR) | | |
| *BRCA2* | $\geq 2^*$ | 48 | 202 | 7.25 (STR) | 5.29 (STR) | 189 | 7.16 (STR) |
| | $\geq 1^*$ | 69 | 321 | 7.88 (STR) | 5.78 (STR) | | |
| *PALB2* | $\geq 2^*$ | 1 | 11 | 3.27 (MOD) | 0 (None) | 101 | 6.30 (STR) |
| | $\geq 1^*$ | 2 | 25 | 4.40 (STR) | 0.95 (None) | | |
| *ATM* | $\geq 1^*$ | 39 | 26 | 4.45 (STR) | 5.00 (STR) | 149 | 6.83 (STR) |
| | $\geq 2^*$ | 59 | 52 | 5.37 (STR) | 5.57 (STR) | | |
| *CHEK2* | $\geq 2^*$ | 4 | 1 | 0 (None) | 1.89 (SUP) | 12 | 3.39 (MOD) |
| | $\geq 1^*$ | 6 | 5 | 2.20 (MOD) | 2.45 (MOD) | | |
| *RAD51C* | $\geq 1^*$ | 7 | 2 | 0.95 (None) | 2.66 (MOD) | 31 | 4.69 (STR) |
| | $\geq 2^*$ | 7 | 4 | 1.89 (SUP) | 2.66 (MOD) | | |
| *RAD51D* | $\geq 2^*$ | 0* | 3 | 1.50 (SUP) | - | 33 | 4.77 (STR) |
| | $\geq 1^*$ | 0* | 4 | 1.89 (SUP) | - | | |
| *BRIP1* | $\geq 1^*$ | 3 | 6 | 2.45 (MOD) | 1.50 (SUP) | 43 | 5.14 (STR) |
| | $\geq 2^*$ | 6 | 25 | 4.40 (STR) | 2.45 (MOD) | | |

We then explored a model of benignity in which a single benign evidence item of any strength (including a single item of supporting evidence) was sufficient for truthset assignation (Table S1). Using this approach, and requiring ≥2/≥5 variant carriers for BS1/BA1, we observed that a much greater number of missense variants were eligible for truthset inclusion, ranging from 1026 (for *BRCA1*) to 14,342 (for *BRCA2*). This then meant that for all genes the size of benign missense truthset would be readily sufficient to generate PS3 strengths of very strong (applying assumption of perfect concordance). However, assigning benignity (B/LB) status on a lesser threshold of evidence is likely to incorporate larger numbers of “false positive” assignations of benignity.

### Improvement upon and correlation with existing clinical classifications

To evaluate the concordance of our proactive-systematic approach with existing clinical classifications, we compared the specific variants comprising benign missense truthsets generated using our “standard” proactive-systematic construction methodology (as defined above, and requiring ≥2/≥5 variant carriers for application of BS1/BA1 population frequency codes) with those generated using existing ClinVar ≥2* B/LB classifications (Table 4; Figure S5). Notably, for four of the surveyed HBOC genes (*PALB2*, *CHEK2*, *RAD51C* and *BRIP1*), our proactive-systematically generated benign missense truthsets included all variants present in the respective ClinVar-derived truthset (*n* = 11, 1, 2, and 5, respectively). For *ATM*, five ≥2* B/LB ClinVar variants were not captured by our proactive-systematic approach; for *BRCA1* and *BRCA2*, there were 157 and 118 such variants, respectively.

As noted above, our approach implements a filter precluding any variant with an existing ClinVar classification of pathogenic (≥1*) from inclusion in a benign truthset. However, we noted that this filtering step did not trigger exclusion of any variants from “standard” proactive-systematically generated benign variant truthsets for any of the surveyed genes, including *BRCA1* and *BRCA2*, which have substantial numbers of missense variants with ≥1* pathogenic ClinVar classifications (172 and 69, respectively). This suggests our approach is unlikely to falsely ascribe benignity to pathogenic variants.

## Discussion

Pathogenic variant classifications are typically deemed of greater clinical value to perform and deposit. By this corollary, counts of available benign-classified variants would be anticipated to lag behind on account of lower motivation for classification and deposition into variant databases by clinical diagnostic laboratories. However, as we demonstrate, there is a paradoxical relationship for clinical assay validation, namely that it is the number of benign-classified variants available for truthsets that most strongly determines the evidence strength towards pathogenicity applicable following clinical validation of a new functional assay. We also demonstrate the paucity of benign-classified missense variants available for the majority of 116 clinically tested cancer susceptibility genes, which is the case even for well-studied, frequently tested HBOC genes. We demonstrate even greater paucity when restricting to those missense variants classified firmly as benign (rather than likely benign) and/or with 2* review status (i.e. more than just a single submitter).

In this study, we present a novel proactive-systematic approach for construction of benign missense variant truthsets that we apply to eight HBOC genes. This methodology could readily be extended to other genes: primarily limiting to this is calculation of the estimated BA1/BS1/BS1_sup thresholds via calculation of an MTAF, which requires clinical expertise in the gene-phenotype dyad (and appraisal of often-complex literature to attain estimates for phenotype/disease incidence, penetrance and/or attribution).

We demonstrate the substantial uplift afforded by our proactive-systematic approach for six of eight HBOC genes widely tested clinically. *BRCA1*/*BRCA2* are outliers in part because of the markedly longer timecourse and higher volume of clinical testing of *BRCA1*/*BRCA2* compared to the other genes. Also influential is the early inception of the ENIGMA group who have instigated frameworks and variant classification activity, long predating the wider establishment of VCEPs.^31^ Most impactfully, landmark multifactorial genetic epidemiologic analyses published in 2007 leveraged unique *BRCA1/BRCA2* clinical datasets assembled from Myriad Genetics, who at that time held the international patent for diagnostic analysis of *BRCA1* and *BRCA2.* ^32^ These and downstream multifactorial-type classifications utilised (i) family history scoring, (ii) co-occurrence in trans with a known pathogenic variant and (iii) co-segregation with disease in pedigrees to generate large numbers of classifications but did not include either population frequency or *in silico* predictions, thus consistent with the lower degree of overlap for these variants with our proactive-systematically generated set.^33–36^.

Also of note is the paucity of *BRCA1* missense variants meeting the criteria for benignity discrepancy during proactive-systematic truthset generation. This is a consequence of the distribution of REVEL scores for *BRCA1* missense variants being skewed towards the deleterious range, thereby precluding the majority from attaining BP4 at any strength.

### Limitations

We have sought in our proactive-systematic benignity classification approach to incorporate accessible data for filtering out variants with indicators of possible pathogenicity (namely *in silico* predictions suggesting protein effect or splicing impact, indicative case-control signal and/or suggestive ClinVar classifications). However, this approach does not identify/exclude existence of every type of evidence indicative of potential pathogenicity that might/should have been sought as part of a manual, bespoke clinical classification. For example, albeit highly infrequent, reports in the literature of the variant identified in biallelic configuration in cases of Fanconi anaemia.

At the time of developing the proactive-systematic benignity classification approach, we continue to be in transition from the 2015 v3.0 ACMG/AMP Richards *et al.* framework (and various evolutions thereof by VCEPs), to the piloting of the draft v4.0 ClinGen/ACMG/AMP Framework, with accordant disparity and ambiguity in best practice for application of evidence codes, weightings and thresholds. For example, in the v3.0 2015 ACMG/AMP framework there are only two levels delineated for population codes (BA1 and BS1). As per the evolution of this by a number of the VCEPs^26–29^, we have employed three strengths of benignity scoring for population frequency: BA1, BS1 and BS1_supporting. Notably, there are likewise three levels described for benignity scoring for population frequency in the in the draft version of the v4.0 ACMG/AMP framework^30^.

Furthermore, and as previously discussed, whilst published thresholds for REVEL and BayesDel were validated against a large dataset^24^, *in silico* prediction tools are recognised to perform differentially for different genes. This could be obviated by allowing multiple in silico tools within our proactive-systematic benignity classification approach, albeit this introduces additional permissiveness.

### Additional challenges in generation of truthsets for clinical validation of MAVES

In assessing the power (evidence strength) afforded from a given number of truthset classifications available for clinical validation of a hypothetical MAVE, we assume (i) there is an assay readout for every truthset variant and (ii) perfect concordance between assay readouts and truthset-classifications (i.e. a perfectly discriminatory assay). In practice, neither of these standards is likely to be attained for a real assay: an assay will likely miss readouts for some of the variants present in the truthset and there will likely be some level of discordance. What we present are thus “best-case scenarios”.

Another challenge may be domain bias: *in silico* tools may more easily classify variants as benign in non-functional domains compared to functional domains. Assays, in particular non-saturation MAVEs, may only target specific regions of the gene of greater interest, namely the functional domains. Accordingly, the expansion of benign missense truthsets will likely disproportionately better serve MAVEs adopting a saturation approach and/or MAVEs that include non-functional domains.

There will also be considerable variation gene-by-gene in factors influencing the maximum size of benign missense truthset that can be generated even using the proactive-systematic approach, which include gene length (and, relatedly, the number of possible missense variants) and general constraint against missense variation.

We demonstrate the sizeable uplift in benignity classifications attainable when applying a more permissive evidence requirement for attaining classification as LB (≥1 evidence item of supporting strength), reflecting the proposed threshold in the v4.0 ACMG/AMP Framework^30^. It is likely overall advantageous to enhance variant numbers (power) at the expense of potential minor contamination with “false positives”.

The same considerations apply when mandating a number of observed variant carriers to attain evidence for population frequency. We elected to mandate ≥2/≥5 variant carriers for BS1/BA1 in our proactive-systematic approach. This prevented inclusion of variants whose benignity was predicated on a single, possibly stochastic, observation. Enforcing a higher variant carrier threshold (e.g. ≥5 throughput) would be expected to increase the precision of population frequency estimates but be punitive to attaining BS1/BS1_sup from smaller population datasets.

Whilst for most genes the number of B/LB missense variant classifications is particularly low, the number of P/LP missense variant classifications is likely also limiting. More systematic approaches to construction of pathogenic truthsets are thus also urgently required: these will likely be predicated on the challenging endeavour of assembly and integration of large clinical datasets.

Genes in which different variant types are associated with multiple disparate disease phenotypes require additional caution. Clinical validation of the assay should be undertaken for each disease phenotype separately, with appropriate generation of benign and pathogenic missense variant sets.

Related to this, we have thus far applied the assumption that all missense variants throughout the gene will be acting by an identical mechanism to the selected missense pathogenic variant truthset; in practice, missense variants in different domains may plausibly have different mechanisms of action related to disease. Whilst there is expert consensus that best practice would be to have domain-specific validation of assays^15^, not only does this further exacerbate the problem of truthset paucity, but the definition of domains is also not straightforward.

A further consideration is the likelihood that many variants, and in particular missense variants, will have effect sizes of intermediate magnitude (reduced penetrance). Whilst in concept assays might present a potential solution for providing a readout to quantify more sensitively disease effect beyond a dichotomous assignation of pathogenic/benign, to provide such a “quantitative” validation of the assays is an even more massive challenge. This would require truthsets of not just of established pathogenicity, but for which variants had been accurately quantified by orthogonal approaches as being of reduced penetrance.

In conclusion, whilst MAVEs provide a highly promising approach by which to advance classification of rare variants newly identified on clinical testing, the paucity of available truthset classifications is a substantive roadblock to the robust clinical validation required for clinical adoption of these new assays. Urgently required are proactive systematic approaches by which available data are used to generate variant classifications, such as that we present here to generate truthsets of benign missense variants.

## Data Availability

All data produced in the present work are contained in the manuscript except Table S2 (available upon reasonable request to the authors)

## Acknowledgements

A.G. and H.H. are supported by Cancer Research UK Catalyst award CanGene-CanVar [C61296/A27223]. S.A. and C.F.R. are supported by CG-MAVE, Cancer Research UK Programme Award [EDDPGM-Nov22/100004].

## Author contributions

C.T. devised the initial proactive-systematic approach and all authors contributed to its evolution. C.F.R. and S.A. conducted all analyses and generated all figures for use in the manuscript. C.T. and C.F.R. produced the initial manuscript draft and all authors were involved in proofreading of this and all subsequent drafts of the manuscript.

## Data and code availability

The published article includes all datasets generated or analyzed during this study.

