## Supplemental Material for "Availability of benign missense variant “truthsets” for validation of functional assays: current status and a novel systematic approach"

### Supplemental Data

#### Supplemental Methods

##### *Extraction of existing ClinVar missense variant classifications in 116 inherited cancer susceptibility genes*

All existing ClinVar classifications for missense variants with review status ≥1* and/or ≥2* were collated and tallied for 116 established hereditary cancer susceptibility genes (CSGs; Figure S2) using the ClinVar flat file dated 21 April 2024. Quantification of applicable evidence strength for a hypothetical MAVE was carried out as previously described[1] for each gene using each of the respective sets of ≥1* and ≥2* ClinVar pathogenic and benign missense variants as truthsets.

This approach assumed (i) all truthset variants were evaluated by the MAVE, (ii) the MAVE exhibited perfect discrimination between variants, (iii) existing functional evidence had not contributed to the ClinVar classifications of truthset variants, and (iv) all ClinVar annotations correctly reflected true pathogenicity or benignity.

Using the MaveRegistry database[2], we further annotated each CSG as being the target of an existing published MAVE, the target of an in-process MAVE, or having no record of an existing or in-process MAVE.

##### *Categorisation of ClinVar comments for existing benign missense variant classifications*

For all eight HBOC genes of interest, the ClinVar database was queried via the API to retrieve all comments attached to benign germline classification submissions for all missense variants. Each comment was assigned one of five classes indicating the level of contribution of functional evidence:

**Functional evidence applied:** Comment explicitly indicates the application of evidence from an in-house or previously published functional study, or mentions the application of the BS3 evidence code.

**Functional evidence explicitly not applied:** Comment explicitly states that functional evidence was not applied or was unavailable for the variant.

**Functional evidence implicitly not applied:** Comment delineates applied evidence items but does not make mention of functional evidence.

**Functional evidence application ambiguous:** Contribution of functional evidence to classification is unclear, e.g. mention of “impact on protein function” without reference to a specific publication or in-house assay.

**No comment:** No text comment was provided with the germline classification submission.

The per-submission functional evidence contribution annotations for a given variant were collapsed to provide a per-variant annotation of functional evidence contribution. This collapsed annotation corresponded to that of the submission with the functional evidence annotation highest in the following hierarchy (from highest to lowest precedence): functional evidence explicitly not applied; functional evidence implicitly not applied; functional evidence applied; functional evidence application ambiguous; no comment. The resulting collapsed analysis is presented in Figure S3, below.

##### *Selection of transcripts of interest*

For each of our hereditary breast and ovarian cancer (HBOC) susceptibility genes of interest, the corresponding MANE transcript was selected for downstream annotation of variant consequence. Bespoke Python scripts were used to generate all possible SNVs in the coding regions of the respective MANE transcripts and the resulting variant list filtered to retain only those resulting in a missense consequence. BC, breast cancer; OC, ovarian cancer.

| **Gene** | **Associated cancer type** | **RefSeq transcript** |
|---|---|---|
| *BRCA1* | Breast | NM_007294.3 |
| *BRCA2* | Breast | NM_000059.3 |
| *PALB2* | Breast | NM_024675.3 |
| *ATM* | Breast | NM_000051.3 |
| *CHEK2* | Breast | NM_007194.3 |
| *RAD51C* | Ovarian | NM_058216.1 |
| *RAD51D* | Ovarian | NM_002878.3 |
| *BRIP1* | Ovarian | NM_032043.3 |

*Definition of UK Biobank breast and ovarian cancer cohorts*

UK Biobank (UKB) samples were first filtered to retain only individuals with a self-reported (data field p31) and genetic (p22011) sex of "Female" and no evidence of aneuploidy (p22019). We additionally excluded any individuals who had previously withdrawn from the UKB cohort.

Registry-linked ICD10 (p40006) and ICD9 (p40013) codes pertaining to personal history of cancer were extracted for all remaining participants. Individuals with any record of any of the below ICD10/9 codes pertaining to personal history of breast and/or ovarian cancer were assigned to our breast and/or ovarian cancer case cohorts, respectively.

| **Category** | **ICD10** | **ICD9** |
|---|---|---|
| *Breast cancer* | | |
| Malignant neoplasm of breast | C50.X | 174X |
| Carcinoma *in situ* of breast | D05.X | 2330 |
| Neoplasm of uncertain behaviour of breast | D48.6 | 2383 |
| *Ovarian cancer* | | |
| Malignant neoplasm of ovary | C56.X | 1830 |
| Benign neoplasm of ovary | D27.X | 220 |
| Neoplasm of uncertain behaviour of ovary | D39.1 | 2362 |

We additionally assigned to the respective case cohorts any individual with a self-reported cancer code (p20001) of "breast cancer" or "ovarian cancer". All individuals not present in the breast cancer or ovarian cancer case cohorts were assigned to the "non-breast cancer" and "non-ovarian cancer" control cohorts, respectively.

Based on breast cancer status, we further filtered participants to exclude related individuals, excluding one of each related pair (genetic relatedness factor >0.17; data field p22011). Where pairs comprised a case and control, the participant in the case cohort was preferentially retained in the final cohort; where both individuals were in the same cohort, one individual in each pair was randomly excluded.

*Assignation of collapsed UKB participant ethnicities*

Self-reported ethnicity fields (p21000) were extracted for all individuals in UKB passing the above cohort filtering steps. To allow downstream stratification of case-control signal and population frequency estimates, each individual was assigned a "collapsed" ethnicity equating to the high-level parent term of their self-reported ethnicity (one of "White", "Black", "Asian", "Chinese", "Mixed" and "Other"; see below). Where multiple self-reported ethnicities were listed for a single participant, a collapsed participant ethnicity was only generated if all listed ethnicities belonged to the same parent term; otherwise, participants were nominally assigned to the "Other" cohort.

| Collapsed ("parent") ethnicity | Granular self-reported ethnicity (data field p21000) |
|---|---|
| **White** | White* |
| | British |
| | Irish |
| | Any other white background* |
| **Mixed*** | Mixed* |
| | White and Black Caribbean* |
| | White and Black African* |
| | White and Asian* |
| | Any other mixed background* |
| **Asian or Asian British** | Asian or Asian British* |
| | Indian |
| | Pakistani |
| | Bangladeshi |
| | Any other Asian background* |
| **Black or Black British** | Black or Black British* |
| | Caribbean |
| | African |
| | Any other Black background* |
| **Chinese** | Chinese |
| **Other*** | Other ethnic group* |
| | Do not know* |
| | Prefer not to answer* |

Collapsed and granular ethnicities marked with asterisks (*) were considered uninformative for population frequency analysis (see "Application of population frequency codes (BA1, BS1 and BS1_sup) for missense variants observed in gnomAD and UK Biobank", below).

*Processing of UK Biobank data and generation of variant counts from gnomAD and UK Biobank*

Variants overlapping the coding sequence of selected HBOC transcripts were extracted from the corresponding UKB population VCFs (pVCFs) using the bcftools view command. As a broad QC step, we filtered variants to retain only those for which there was total read coverage >10 at the relevant site in >90% of total UKB samples. Bespoke scripts were then used to tally the total number of heterozygous and homozygous carriers of each variant. Allele counts were additionally stratified according to the collapsed ethnicity of carriers, as defined above, as well as according to their case/control status.

For gnomAD, counts of variants overlapping the target regions of the selected HBOC transcripts in female samples were extracted from the v2.1.1 flat file download, again using bcftools view. This included both total variant counts and ancestry-stratified counts.

All variants identified in one or both datasets were annotated with their consequence against the corresponding MANE transcript using the Ensembl Variant Effect Predictor v111 and then filtered to retain only those with a "Consequence" field containing the string "missense_variant".

The resulting variant set (and associated counts) were merged with those published in the missense portion of the BRIDGES study[3] to produce a finalised missense variant set comprising any variant identified in either study, alongside the associated variant counts.

*Calculation of maximum tolerated allele frequencies (MTAFs) for eight HBOC genes*

For application of the BA1/BS1/BS1_sup population codes, we applied the approach detailed by Whiffin et al.[4] and widely incorporated into VCEP guidance across multiple diseases for generation of a maximum tolerated allele frequency (MTAF). The MTAF reflects the population frequency above which a variant would not reasonably be expected to exist given disease penetrance, prevalence and genetic/allelic heterogeneity, and can be readily applied as a BA1 allele frequency threshold. Estimates of genetic heterogeneity were derived from empirical studies of mutational frequency in unselected breast[3] (*BRCA1*, *BRCA2*, *PALB2*, *ATM*, *CHEK2*) and ovarian[5] (*RAD51C*, *RAD51D* and *BRIP1*) cancer patients. Where available (all genes except *ATM* and *CHEK2*), penetrance estimates were taken from existing UKCGG clinical guidelines and their source studies[6-9]; the estimates for *ATM* and *CHEK2* were taken from published penetrance analyses[10,11]. The distinct BA1, BS1 and BS1_sup allele frequency thresholds were calculated by specifying allelic heterogeneity values of 1, 0.1 and 0.05 during MTAF construction, respectively.

MTAFs and corresponding population frequency thresholds were constructed using the following parameters:

| Gene | Associated phenotype | Prevalence (1 in...) | Penetrance | Genetic heterogeneity | BA1 | BS1 | BS1_sup |
|---|---|---|---|---|---|---|---|
| ***BRCA1*** | BC | 8 | 0.72 | 0.011 | $9.16x10^{-4}$ | $9.16x10^{-5}$ | $4.58x10^{-5}$ |
| ***BRCA2*** | BC | 8 | 0.69 | 0.015 | $1.40x10^{-3}$ | $1.40x10^{-4}$ | $6.99x10^{-4}$ |
| ***PALB2*** | BC | 8 | 0.53 | 0.0056 | $6.62x10^{-4}$ | $6.62x10^{-5}$ | $3.31x10^{-5}$ |
| ***ATM*** | BC | 8 | 0.33 | 0.0060 | $1.14x10^{-3}$ | $1.14x10^{-4}$ | $5.70x10^{-4}$ |
| ***CHEK2*** | BC | 8 | 0.24 | 0.014 | $3.75x10^{-3}$ | $3.75x10^{-4}$ | $1.88x10^{-4}$ |
| ***RAD51C*** | OC | 50 | 0.11 | 0.0057 | $5.22x10^{-4}$ | $5.22x10^{-5}$ | $2.61x10^{-5}$ |
| ***RAD51D*** | OC | 50 | 0.13 | 0.0057 | $4.42x10^{-4}$ | $4.42x10^{-5}$ | $2.21x10^{-5}$ |
| ***BRIP1*** | OC | 50 | 0.10 | 0.014 | $1.36x10^{-3}$ | $1.36x10^{-4}$ | $6.79x10^{-5}$ |

*Application of population frequency codes (BA1, BS1 and BS1_sup) for missense variants observed in gnomAD and UK Biobank*

For accurate application of population frequency codes, we first identified a set of informative non-founder and non-admixed ancestries. This comprised, from gnomAD, all ancestry groupings except “Admixed American”, “Ashkenazi Jewish”, “European (Finnish)” and “Remaining individuals”. For UKB, we considered all collapsed ancestries except “Mixed” and “Other”, and additionally included participants’ granular subethnicities in downstream analysis, with the exception of uninformative ethnicities indicated with an asterisk (*) in the table in “*Assignation of collapsed UKB participant ethnicities*”, above.

Evidence of high population frequency was evaluated for all missense variants present in one or both of gnomAD and UKB by considering each informative ancestry and (for UKB) granular subethnicity in turn, as well as the total set of participants in each dataset. As described in Whiffin et al.[4], we calculated a maximum tolerated allele count (MTAC) by first modelling the variant counts in the ancestry grouping (or total participant set) as a Poisson distribution in which the number of events equals the total number of samples in that ancestry and the average rate of success equals the MTAF. The MTAC is generated using the inverse cumulative distribution function of the Poisson distribution via the qpois function in R, setting the percentile to 0.95 and the lambda value to the MTAF multiplied by the total allele number, i.e. the expected number of successes. This generates a value equivalent to the observed allele count expected to correspond, with a confidence level of 95%, to a true underlying allele frequency meeting or exceeding the corresponding MTAF in that ancestry group.

BA1, BS1 and/or BS1_sup codes were assigned to a variant where the observed number of variant carriers exceeded the respective MTAC in any collapsed ethnicity grouping or granular subethnicity across all participants. We additionally enforced hard thresholds of required carriers alongside the ancestry- and gene-specific MTAC threshold, variously requiring ≥1, ≥2 or ≥5 carriers for application of the population frequency codes to investigate the impact of varying stringency on the numbers of ascertained truthset variants.

We manually excluded three variants for which allele frequency in the population datasets was >50% and therefore indicative of the reference allele being the minor allele. These were *ATM* c.5948A>G (all sequenced alleles in gnomAD; AF=100%), *BRCA2* c.7397T>C (504,076/504,974 sequenced alleles in UKB; AF=99.8%) and *BRIP1* c.2755T>C (302,627/505,290 sequenced alleles in UKB; AF=59.9%). These variants were also excluded from any ClinVar-derived truthsets.

*Exclusion of missense variants with evidence of case-control signal in BRIDGES and UK Biobank sequencing datasets*

As an additional filter against inclusion of potentially pathogenic variants in our proactive-systematically classified truthsets, we quantified case-control signal towards breast/ovarian cancer for missense variants identified in (i) the BRIDGES breast cancer population datasets[3] (breast cancer genes only) and (ii) UKB (breast and ovarian cancer genes). Counts were stratified according to the number of case and non-case carriers based on the gene-associated phenotype (breast or ovarian cancer), with BRIDGES breast cancer case cohorts being as listed in the downloaded datasets, UKB breast and ovarian cancer case cohorts being derived via the cohorting approach described above.

Odds ratios (ORs), 95% confidence intervals (CIs) and p-values (based on the Fisher exact test) were calculated based on observed counts of carriers and non-carriers between cases and controls independently in BRIDGES and UKB for each variant. Variants were excluded from benign truthsets where the OR exceeded 2 with a lower CI limit >1 in one or both datasets.

#### Supplemental Figures

**Figure S1.** *Workflow for proactive-systematic generation of benign missense truthsets.* **(A)** Conventional truthset construction may often rely on classifications deposited in ClinVar. Our proactive-systematic approach instead uses 2015 ACMG/AMP-informed code combinations; namely, either the annotation of a variant with a BA1 flag, or with both a BS1/BS1_sup population frequency flag and a BP4 *in silico* flag of any strength. **(B)** The proactive-systematic approach to benign missense truthset construction begins with the *in silico* generation of all possible SNVs in a transcript of interest. Variants are precluded from inclusion in benign truthsets if annotated as pathogenic or likely pathogenic with ≥1* review status in ClinVar. Any variant with evidence of case-control signal of odds ratio (OR) >2 with lower confidence interval (CI) >1 in a suitable sequencing dataset. Variants with any *in silico evidence* of pathogenicity in terms of BayesDel (≥0.13), REVEL (≥0.644) and SpliceAI (≥0.2) scores are similarly excluded from truthset inclusion, while BP4 flags indicating benignity are assigned according to previously calibrated REVEL thresholds[12]. Population codes indicative of benignity are then applied according to gene-specific thresholds, as described in the Methods.

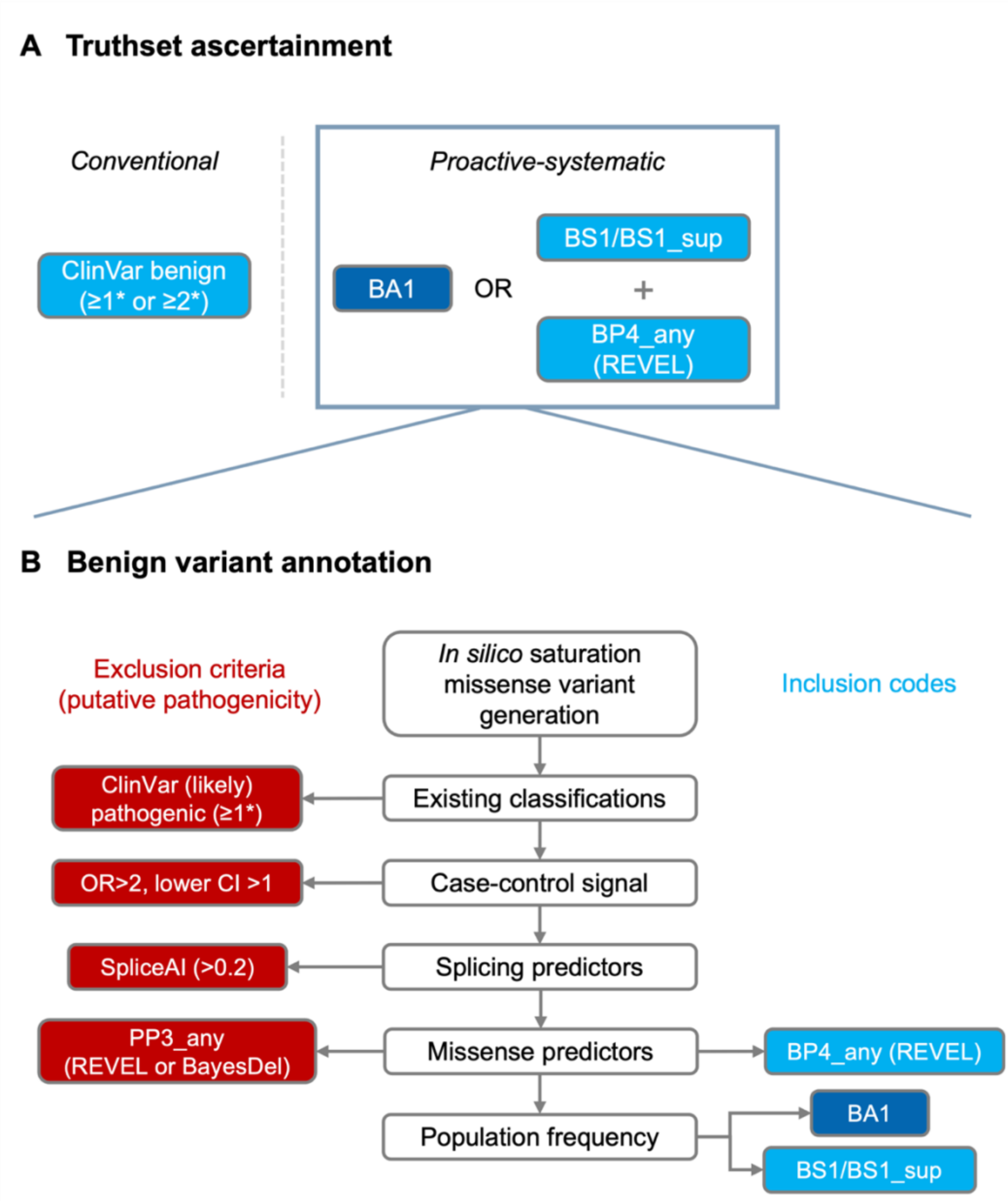

**Figure S2.** *Hypothetical applicable PS3 evidence strengths for MAVEs targeting 116 inherited cancer susceptibility genes if using ClinVar-derived benign missense truthsets.* Existing (likely) benign missense variant classifications with **(A)** ≥1* and **(B)** ≥2* review status were collated from ClinVar and used as the hypothetical benign truthsets for perfectly concordant MAVEs. The resulting counts were converted to the maximum applicable PS3 exponent points (left) and evidence strengths (right) for truthsets of the respective size under the assumption that there was at least one variant in the pathogenic truthset, as PS3 strength can be calculated using benign truthset size alone for perfectly concordant assays. Where no benign missense variants were present in ClinVar, a PS3 strength could not be calculated and the resulting $EPs_{Path}$ was designated "NA". CSG, cancer susceptibility gene; $EPs_{Path}$, exponent points towards pathogenicity; MOD, moderate; STR, strong; SUP, supporting.

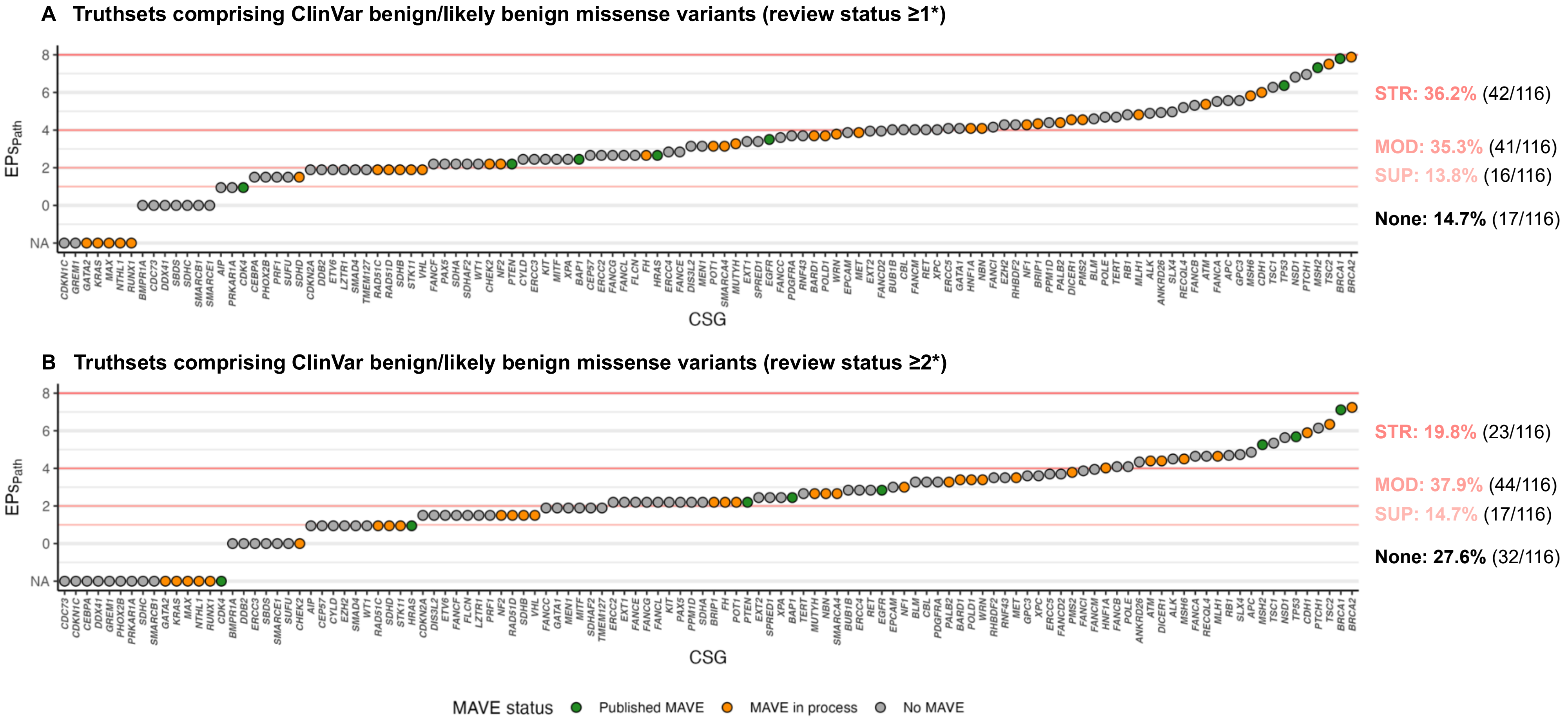

**Figure S3.** *Summary of inclusion of functional data in ClinVar classifications for missense variants in eight HBOC genes.* All available ClinVar classifications and their associated comments were extracted for all missense variants with classifications of benignity in eight hereditary breast and ovarian cancer (HBOC) genes of clinical interest. A single classification class was assigned to each variant based on the presence of any classification for that variant. Absence of functional data was deemed explicit if the comment mentioned a lack of available functional data, and implicit if other evidence items were listed with no mention of functional evidence.

**A All benign missense ClinVar classifications (≥1*)**

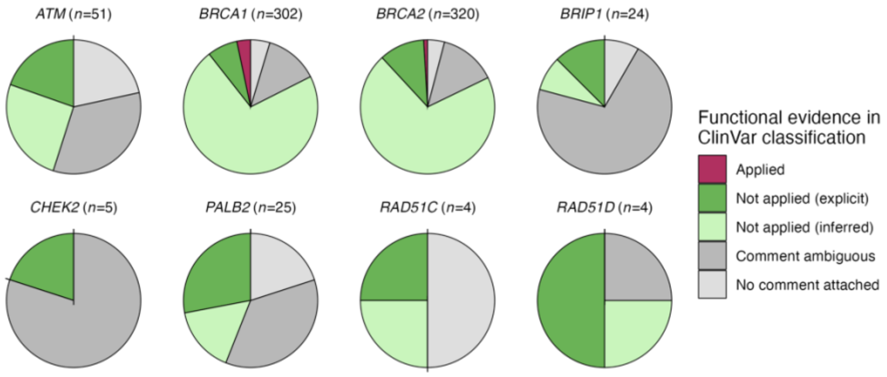

**B 1* benign missense ClinVar classifications**

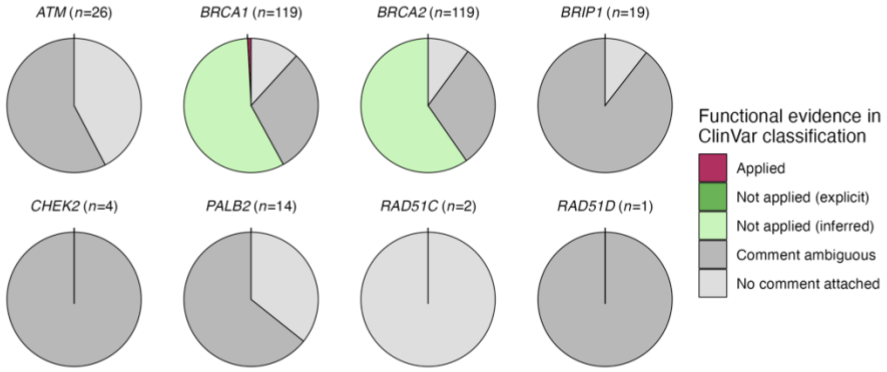

**C 2* benign missense ClinVar classifications**

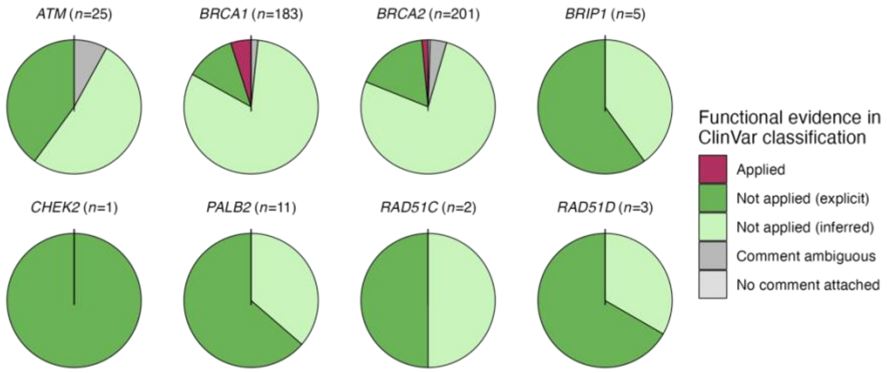

**Figure S4.** *Relative contributing allele counts by combination of evidence items applied in the proactive-systematic approach to benign missense truthset construction.* **(A)** We observed that, as expected, increasingly stringent population evidence codes were associated with higher allele counts in the contributing populations, ranging from 2-77 for BS1_sup, 2-585 for BS1 and 5-177,212. **(B)** Considering low-frequency variants (<10 allele count) in isolation, we observed that only for four variants in PALB2 is an allele count of 5 (for one variant) or 6 (for three variants) sufficient to attain BA1; these variants are present in the small UKB African and Chinese sub-ancestries. Excluding PALB2 variants, the range of allele counts permitting application of BA1 changes to 10-177,212.

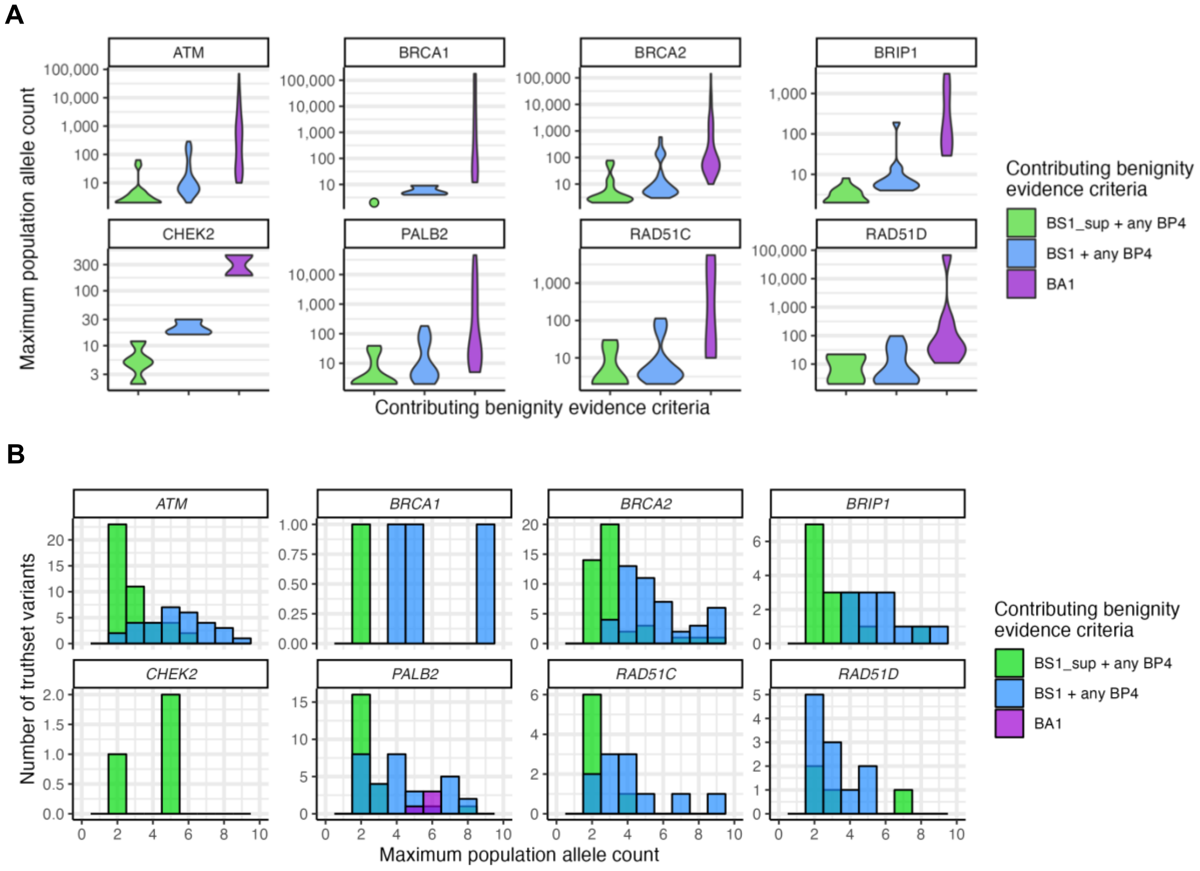

**Figure S5.** *Concordance between variants comprising benign missense truthsets generated via ClinVar and our proactive-systematic approach.* Benign missense truthsets were constructed using ClinVar-annotated variants with review status ≥2* and using our proactive-systematic ACMG/AMP-based approach (as defined in main text), requiring ≥2 variant carriers for assignation of population frequency codes. Shown are the counts of and overlap between variants assigned to truthsets using each methodology.

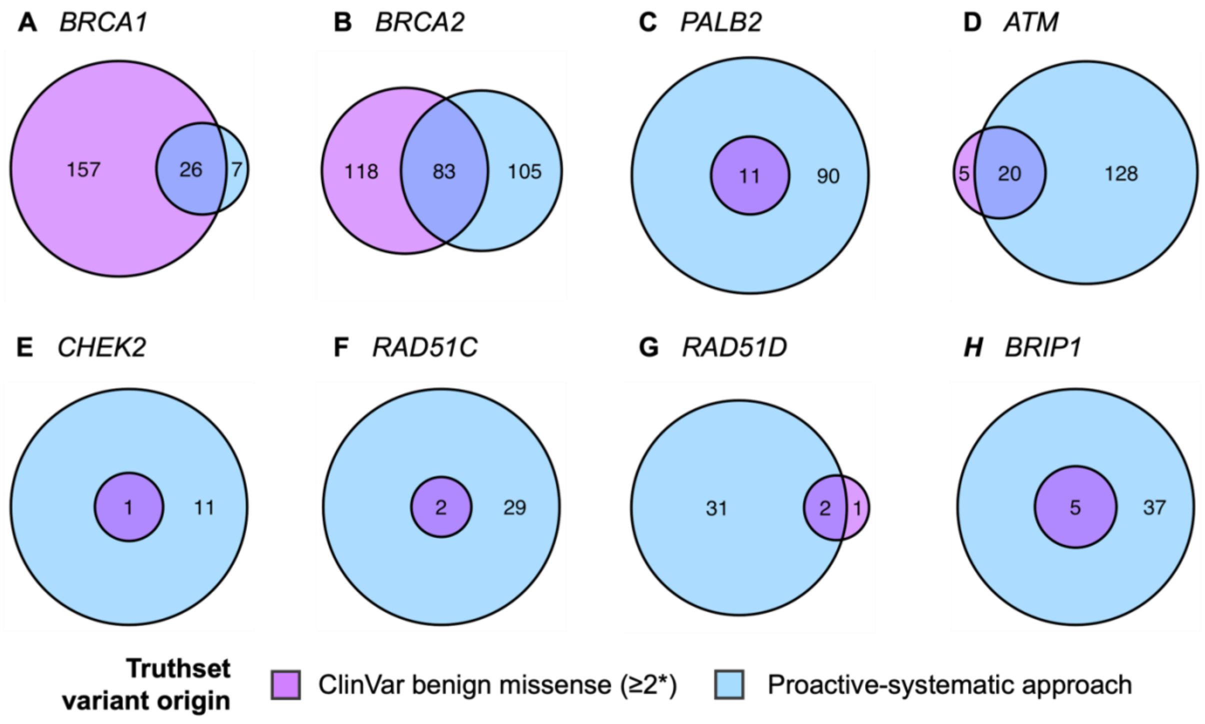

#### Supplemental Tables

**Table S1.** *Number of proactive-systematically generated benign truthset variants and corresponding MAVE strengths for eight HBOC genes.* Displayed are the number of variants assigned to benign truthsets using both conventional ClinVar-based approaches (variants with ≥2* and ≥1* review status), and various formulations of proactive-systematic construction, as detailed in the main text*. With the exception of the ClinVar ≥2* approach, pathogenic truthsets comprised all pathogenic variants with ClinVar review status ≥1*. Highlighted in grey are counts for the standard approach of ACMG evidence combinations and requiring ≥2 variant carriers for BS1, BS1_sup and ≥5 variant carriers for BA1, the approach selected for presentation in the main text.
EPs, exponent points. Required variant carriers: ≥1= ≥1variant carrier sufficient for any population codes; ≥2/≥5 = ≥2 variant carriers required for BS1/BS1_sup, ≥5 variant carriers required for BA1; ≥5 = ≥5 variant carriers required for any population code.
* For *RAD51D*, an arbitrary pathogenic truthset size of 1 was used for calculation of PS3 EPs and strength due to an absence of eligible pathogenic missense variants in ClinVar.

| Truthset construction methodology | Required variant carriers | Number of missense truthset variants | | Applicable EPs | | Applicable 2015 ACMG strength | |
|---|---|---|---|---|---|---|---|
| | | Pathogenic | Benign | PS3 | BS3 | PS3 | BS3 |
| *BRCA1* (associated phenotype: breast cancer) | | | | | | | |
| ClinVar ≥2* | - | 108 | 183 | 7.11 | 6.39 | STR | STR |
| ClinVar ≥1* | - | 172 | 302 | 7.80 | 7.03 | STR | STR |
| Standard (BA1 or BS1/BS1_sup + any BP4) | ≥1 | 108 | 33 | 4.77 | 6.39 | STR | STR |
| | ≥2/≥5 | 108 | 33 | 4.77 | 6.39 | STR | STR |
| | ≥5 | 108 | 31 | 4.69 | 6.39 | STR | STR |
| Standard + any strong | ≥1 | 108 | 91 | 6.16 | 6.39 | STR | STR |
| | ≥2/≥5 | 108 | 90 | 6.14 | 6.39 | STR | STR |
| | ≥5 | 108 | 68 | 5.76 | 6.39 | STR | STR |
| Standard + any supporting | ≥1 | 108 | 1027 | 9.47 | 6.39 | VSTR | STR |
| | ≥2/≥5 | 108 | 1026 | 9.47 | 6.39 | VSTR | STR |
| | ≥5 | 108 | 986 | 9.41 | 6.39 | VSTR | STR |
| *BRCA2* (associated phenotype: breast cancer) | | | | | | | |
| ClinVar ≥2* | - | 48 | 201 | 7.24 | 5.29 | STR | STR |
| ClinVar ≥1* | - | 69 | 320 | 7.88 | 5.78 | STR | STR |
| Standard (BA1 or BS1/BS1_sup + any BP4) | ≥1 | 48 | 188 | 7.15 | 5.29 | STR | STR |
| | ≥2/≥5 | 48 | 188 | 7.15 | 5.29 | STR | STR |
| | ≥5 | 48 | 135 | 6.70 | 5.29 | STR | STR |
| Standard + any strong | ≥1 | 48 | 247 | 7.52 | 5.29 | STR | STR |
| | ≥2/≥5 | 48 | 246 | 7.52 | 5.29 | STR | STR |
| | ≥5 | 48 | 187 | 7.14 | 5.29 | STR | STR |
| Standard + any supporting | ≥1 | 48 | 14343 | 13.07 | 5.29 | VSTR | STR |
| | ≥2/≥5 | 48 | 14342 | 13.07 | 5.29 | VSTR | STR |
| | ≥5 | 48 | 14320 | 13.07 | 5.29 | VSTR | STR |
| *PALB2* (associated phenotype: breast cancer) | | | | | | | |
| ClinVar ≥2* | - | 1 | 11 | 3.27 | 0.00 | MOD | None |
| ClinVar ≥1* | - | 2 | 25 | 4.40 | 0.95 | STR | None |
| Standard (BA1 or BS1/BS1_sup + any BP4) | ≥1 | 1 | 107 | 6.38 | 0.00 | STR | None |
| | ≥2/≥5 | 1 | 101 | 6.30 | 0.00 | STR | None |
| | ≥5 | 1 | 61 | 5.61 | 0.00 | STR | None |
| Standard + any strong | ≥1 | 1 | 837 | 9.19 | 0.00 | VSTR | None |
| | ≥2/≥5 | 1 | 832 | 9.18 | 0.00 | VSTR | None |
| | ≥5 | 1 | 797 | 9.12 | 0.00 | VSTR | None |
| Standard + any supporting | ≥1 | 1 | 7102 | 12.11 | 0.00 | VSTR | None |
| | ≥2/≥5 | 1 | 7102 | 12.11 | 0.00 | VSTR | None |
| | ≥5 | 1 | 7099 | 12.11 | 0.00 | VSTR | None |
| *ATM* (associated phenotype: breast cancer) | | | | | | | |
| ClinVar ≥2* | - | 39 | 25 | 4.40 | 5.00 | STR | STR |
| ClinVar ≥1* | - | 59 | 51 | 5.37 | 5.57 | STR | STR |
| | ≥1 | 39 | 149 | 6.83 | 5.00 | STR | STR |

| | | | | | | | |
|---|---|---|---|---|---|---|---|
| Standard (BA1 or BS1/BS1_sup + any BP4) | ≥2/≥5 | 39 | 148 | 6.82 | 5.00 | STR | STR |
| | ≥5 | 39 | 100 | 6.29 | 5.00 | STR | STR |
| Standard + any strong | ≥1 | 39 | 205 | 7.27 | 5.00 | STR | STR |
| | ≥2/≥5 | 39 | 202 | 7.25 | 5.00 | STR | STR |
| | ≥5 | 39 | 150 | 6.84 | 5.00 | STR | STR |
| Standard + any supporting | ≥1 | 39 | 10156 | 12.60 | 5.00 | VSTR | STR |
| | ≥2/≥5 | 39 | 10154 | 12.60 | 5.00 | VSTR | STR |
| | ≥5 | 39 | 10139 | 12.59 | 5.00 | VSTR | STR |
| *CHEK2* (associated phenotype: breast cancer) | | | | | | | |
| ClinVar ≥2* | - | 4 | 1 | 0.00 | 1.89 | None | SUP |
| ClinVar ≥1* | - | 6 | 5 | 2.20 | 2.45 | MOD | MOD |
| Standard (BA1 or BS1/BS1_sup + any BP4) | ≥1 | 4 | 12 | 3.39 | 1.89 | MOD | SUP |
| | ≥2/≥5 | 4 | 12 | 3.39 | 1.89 | MOD | SUP |
| | ≥5 | 4 | 11 | 3.27 | 1.89 | MOD | SUP |
| Standard + any strong | ≥1 | 4 | 28 | 4.55 | 1.89 | STR | SUP |
| | ≥2/≥5 | 4 | 27 | 4.50 | 1.89 | STR | SUP |
| | ≥5 | 4 | 26 | 4.45 | 1.89 | STR | SUP |
| Standard + any supporting | ≥1 | 4 | 1750 | 10.20 | 1.89 | VSTR | SUP |
| | ≥2/≥5 | 4 | 1749 | 10.20 | 1.89 | VSTR | SUP |
| | ≥5 | 4 | 1749 | 10.20 | 1.89 | VSTR | SUP |
| *RAD51C* (associated phenotype: ovarian cancer) | | | | | | | |
| ClinVar ≥2* | - | 7 | 2 | 0.95 | 2.66 | None | MOD |
| ClinVar ≥1* | - | 7 | 4 | 1.89 | 2.66 | SUP | MOD |
| Standard (BA1 or BS1/BS1_sup + any BP4) | ≥1 | 7 | 31 | 4.69 | 2.66 | STR | MOD |
| | ≥2/≥5 | 7 | 31 | 4.69 | 2.66 | STR | MOD |
| | ≥5 | 7 | 18 | 3.95 | 2.66 | MOD | MOD |
| Standard + any strong | ≥1 | 7 | 49 | 5.31 | 2.66 | STR | MOD |
| | ≥2/≥5 | 7 | 49 | 5.31 | 2.66 | STR | MOD |
| | ≥5 | 7 | 35 | 4.85 | 2.66 | STR | MOD |
| Standard + any supporting | ≥1 | 7 | 1552 | 10.03 | 2.66 | VSTR | MOD |
| | ≥2/≥5 | 7 | 1552 | 10.03 | 2.66 | VSTR | MOD |
| | ≥5 | 7 | 1549 | 10.03 | 2.66 | VSTR | MOD |
| *RAD51D* (associated phenotype: ovarian cancer)* | | | | | | | |
| ClinVar ≥2* | - | 0 | 3 | 1.50 | - | SUP | None |
| ClinVar ≥1* | - | 0 | 4 | 1.89 | - | SUP | None |
| Standard (BA1 or BS1/BS1_sup + any BP4) | ≥1 | 0 | 34 | 4.82 | - | STR | None |
| | ≥2/≥5 | 0 | 33 | 4.77 | - | STR | None |
| | ≥5 | 0 | 21 | 4.16 | - | STR | None |
| Standard + any strong | ≥1 | 0 | 81 | 6.00 | - | STR | None |
| | ≥2/≥5 | 0 | 80 | 5.98 | - | STR | None |
| | ≥5 | 0 | 67 | 5.74 | - | STR | None |
| Standard + any supporting | ≥1 | 0 | 1215 | 9.70 | - | VSTR | None |
| | ≥2/≥5 | 0 | 1215 | 9.70 | - | VSTR | None |
| | ≥5 | 0 | 1213 | 9.70 | - | VSTR | None |
| *BRIP1* (associated phenotype: ovarian cancer) | | | | | | | |
| ClinVar ≥2* | - | 3 | 5 | 2.20 | 1.50 | MOD | SUP |
| ClinVar ≥1* | - | 6 | 24 | 4.34 | 2.45 | STR | MOD |
| Standard (BA1 or BS1/BS1_sup + any BP4) | ≥1 | 3 | 42 | 5.10 | 1.50 | STR | SUP |
| | ≥2/≥5 | 3 | 42 | 5.10 | 1.50 | STR | SUP |
| | ≥5 | 3 | 26 | 4.45 | 1.50 | STR | SUP |
| Standard + any strong | ≥1 | 3 | 85 | 6.07 | 1.50 | STR | SUP |
| | ≥2/≥5 | 3 | 85 | 6.07 | 1.50 | STR | SUP |
| | ≥5 | 3 | 68 | 5.76 | 1.50 | STR | SUP |
| Standard + any supporting | ≥1 | 3 | 4554 | 11.50 | 1.50 | VSTR | SUP |
| | ≥2/≥5 | 3 | 4554 | 11.50 | 1.50 | VSTR | SUP |
| | ≥5 | 3 | 4547 | 11.50 | 1.50 | VSTR | SUP |

**Table S2.** *Summary of variants in proactive-systematically generated benign missense truthsets.*

This table is provided as a supplemental Excel spreadsheet alongside the primary manuscript.
